# A validated heart-specific model for splice-disrupting variants in childhood heart disease

**DOI:** 10.1101/2023.11.23.23298903

**Authors:** Robert Lesurf, Jeroen Breckpot, Jade Bouwmeester, Nour Hanafi, Anjali Jain, Yijing Liang, Tanya Papaz, Jane Lougheed, Tapas Mondal, Mahmoud Alsalehi, Luis Altamirano-Diaz, Erwin Oechslin, Enrique Audain, Gregor Dombrowsky, Alex V Postma, Odilia I Woudstra, Berto J Bouma, Marc-Phillip Hitz, Connie R Bezzina, Gillian Blue, David S Winlaw, Seema Mital

## Abstract

Congenital heart disease (CHD) is the most common congenital anomaly. Non-canonical splice-disrupting variants are not routinely evaluated by clinical tests. Algorithms including SpliceAI predict such variants, but are not specific to cardiac-expressed genes. Whole genome (WGS) (n=1083) and myocardial RNA-Sequencing (RNA-Seq) (n=114) of CHD cases was used to identify splice-disrupting variants. Using features of variants confirmed to affect splicing in myocardial RNA, we trained a machine learning model that outperformed SpliceAI for predicting cardiac-specific splice-disrupting variants (AUC 0.92 vs 0.66), and was independently validated in 43 cardiomyopathy probands (AUC 0.88 vs 0.64). Application of this model to 971 CHD WGS samples identified 9% patients with splice-disrupting variants in CHD genes. Forty-one% of predicted splice-disrupting variants were deeply intronic. The burden of variants in CHD genes was higher in cases compared with 2,570 controls. Our model improved genetic yield by identifying splice-disrupting variants that are not evaluated by routine tests.

## INTRODUCTION

Congenital heart disease (CHD) is the most common congenital anomaly, occurring in ∼1% of live births^1^. Children with complex CHD account for a disproportionately high healthcare burden in society. Two common phenotypes of CHD include tetralogy of Fallot (TOF) and dextro-transposition of the great arteries (TGA). Although there is a strong familial and genetic association to CHD^2^, ∼90% of sporadic cases with isolated CHD remain gene-elusive on conventional clinical testing that is typically limited to exons of known disease-associated genes^3–5^.

Among cases where a genetic cause is identified, normal gene function can be disrupted through a variety of mechanisms, including missense variants, premature stop codons, insertions, deletions, or altered RNA splicing. Splice-disruptions may include the loss of wild-type splice junctions and/or the gain of ‘cryptic’ splice sites that create novel exon boundaries, ultimately resulting in disruptions to the normal pattern of RNA splicing which in turn lead to abnormal protein isoforms. Splice-disrupting variants can occur near existing canonical splice sites, in exons, or in deeply intronic regions.

While canonical splice site variants can be identified using conventional sequencing workflows, splice-disrupting variants outside of these sites are more difficult to identify with high confidence. Such non-canonical splice-disrupting variants are reportedly pathogenic in up to 15% of patients with rare genetic disorders^6^ but cannot routinely be evaluated by conventional genetic testing. Recent reports have identified non-canonical splice-disrupting variants in CHD and other rare diseases primarily using *in silico* predictions in whole exome and whole genome sequencing data, followed by *in vitro* validation of their effect using minigene assays^7–9^. However, whole exome sequencing is unable to detect deeply intronic splice-disrupting variants, and minigene assays alone have technical limitations as a patient-relevant functional assay. Further, current models are not specifically designed to identify cardiac specific splice-disrupting variants expressed in the human heart. The use of patient myocardium to identify and validate aberrant splicing events has a strong potential to address this gap^10^. Recent American College of Medical Genetics and Genomics and the Association for Molecular Pathology (ACMG/AMP) framework emphasizes that the effect of splice-disrupting variants can be more accurately validated in patient-derived tissue samples^11^.

Here we used whole genome sequencing (WGS) and myocardial RNA-Sequencing (RNA-Seq) to identify and validate cardiac specific splice disrupting variants and to develop a heart-specific model for canonical and non-canonical splice variants, which can be applied to patients with CHD. These included patients with two of the most common forms of cyanotic CHD i.e. tetralogy of Fallot (TOF) and transposition of the great arteries (TGA). In addition to identifying canonical splice-disrupting variants in known CHD-related genes in 0.5% cases, this approach identified putatively damaging non-canonical splice-disrupting variants in 8% of isolated CHD, with deeply intronic variants representing 41% of all high-confidence splice-disrupting variants in CHD genes. WGS was critical for the identification of variants that would not be captured by routine clinical genetic tests including exome sequencing^12,13^, while cardiac RNA sequencing (RNA-Seq) allowed for high specificity in the interpretation of splice-disrupting effects. This splice-disrupting variant discovery framework, coupled with a case-control burden analysis, provides a practical strategy for increasing the yield of pathogenic variants in known CHD genes.

## RESULTS

### Study cohort

Our overall study included 1085 CHD probands, of which 856 had TOF and 229 had TGA (**Table 1**). Probands with a clinically and/or genetically diagnosed syndrome were excluded. Among these cases, 708 cases were enrolled through the Heart Centre Biobank Registry at the Hospital for Sick Children (Canada), 250 cases were enrolled through the Kids Heart BioBank at the Heart Centre for Children, The Children’s Hospital at Westmead (Australia), and 127 cases were enrolled through the CONCOR registry at the Amsterdam Medical Center (Netherlands). CHD probands were subdivided into a *Discovery* cohort, which included 114 unrelated TOF probands with RNA-Seq data from right ventricular myocardium and WGS data derived from blood or saliva in 112 of the 114 probands, and an *Extension* cohort, which included 971 unrelated isolated CHD probands (742 TOF and 229 TGA) with WGS data. 18% of probands in the *Discovery* and *Extension* cohorts received one or more forms of prior clinical genetic testing, including cytogenetic, microarray, single gene polymerase chain reaction, gene panel, and/or whole exome sequencing, of which 1% of the cohort harbored pathogenic or likely pathogenic protein-coding variants in known CHD-related genes (1% TOF and 0% TGA). An independent cohort of 43 patients with cardiomyopathy in whom WGS and myocardial RNA-seq was available were used for model validation^14^. The collection and use of all biospecimens was approved by local or central Research Ethics Boards, written informed consent was obtained from all patients, parents or legal guardians, and study protocols adhered to the Declaration of Helsinki. 2,570 WGS samples from the Medical Genome Reference Bank^15^ were obtained for use as a Control cohort.

### Protein-coding variants in CHD genes

We first analyzed WGS data to identify pathogenic or likely pathogenic variants in CHD-associated genes (**Table 2**). These included 99 Tier 1 CHD genes with moderate, strong, or definitive associations with CHD according to Clinical Genome Resource (ClinGen) criteria (17 isolated CHD genes, 82 syndromic CHD genes)^16^, and 626 Tier 2 CHD genes with more limited association with CHD. Tier 2 genes were identified using published literature, existing databases including Online Mendelian Inheritance in Man (OMIM)^17^, ClinGen^18^, and CHDgene^19^, their inclusion in clinical gene panels, and expert curation. CHD genes were further annotated by their mode of inheritance and haploinsufficiency intolerance. ACMG/AMP criteria^20,21^ were applied to protein-coding single nucleotide variants (SNVs), insertion-deletions (indels), and copy number variants (CNVs) in CHD genes, yielding pathogenic variants in 5% of probands (5% TOF and 1% TGA) (**Table 1**).

### Discovery of splice-disrupting variants affecting cardiac-expressed genes

We explored WGS data in the *Discovery* cohort for DNA SNVs and indels that were predicted with high sensitivity using SpliceAI^22^ to result in the loss of a wild-type splice junction and/or the gain of a novel cryptic splice site (SpliceAI Δ score ≥ 0.2). This identified 19,448 variants of which 9,610 were rare within the *Discovery* cohort i.e. found in no more than one sample (internal minor allele frequency (MAF) < 0.01). Among these, 427 (4%) occurred at canonical splice sites. Next, we searched for splice-disrupting events in patient myocardium by applying the *in silico* tool FRASER^23^ to myocardial RNA-Seq data which allows detection of not only alternative splicing but also intron retention events^23^. Across the RNA-seq data, 11,251 genes had a TPM expression ≥ 1 i.e. were expressed in cardiac tissue. We limited our selection of splice sites to those where reads from the donor and acceptor sites were aligned within the same gene, and either rarely were observed to be spliced together within the cohort (ψ/θ – Δψ/θ ≤ 0.1) or nearly always were observed to be spliced together (ψ/θ – Δψ/θ ≥ 0.9).

Significant outlier splicing events within a sample were then defined as those having a false discovery rate < 0.2, an absolute Z-score ≥ 1, and an absolute Δψ/θ score ≥ 0.2, indicating that alternative splicing between two sites was observed 20% more or less often than expected. This yielded 695 significantly altered genes having affected splice junctions and/or intron-retention events with a high effect size. A median of 5 genes were affected per sample, and 80 genes were altered in more than one patient.

We then classified the rare DNA variants identified in WGS data by whether or not they were associated with a matching significant outlier splicing event in myocardial RNA-Seq data from the same proband. This yielded 100 DNA variants that were associated with observable splice disruption in the myocardium i.e. confirmed splicing events (**Table 3**), and 9,406 DNA variants where a significant tissue effect was not observed. 104 DNA variants were excluded due to an indeterminate association with an observed splicing event i.e. more than 100bp outside of a significantly altered splice donor/acceptor pair. Thirty-three of the confirmed splicing events occurred at canonical splice sites. An overview of our variant prioritization strategy is shown in **Figure 1**. A comparison of confirmed splice-disrupting variants i.e. DNA variants associated with a splicing event in the myocardium, versus the remaining unconfirmed splice-disrupting variants revealed that true positive variants had higher SpliceAI Δ scores (p=7.5×10^-27^), affected genes with higher RNA expression (p=1.2×10^-28^), were less likely to occur in low complexity regions of the genome (p=0.01), and analogously were more likely to occur in non-repetitive regions of the genome (p=2.8×10^-6^). All confirmed variants were in genes with a median TPM expression > 0.9 in our cohort.

**Figure 1.**
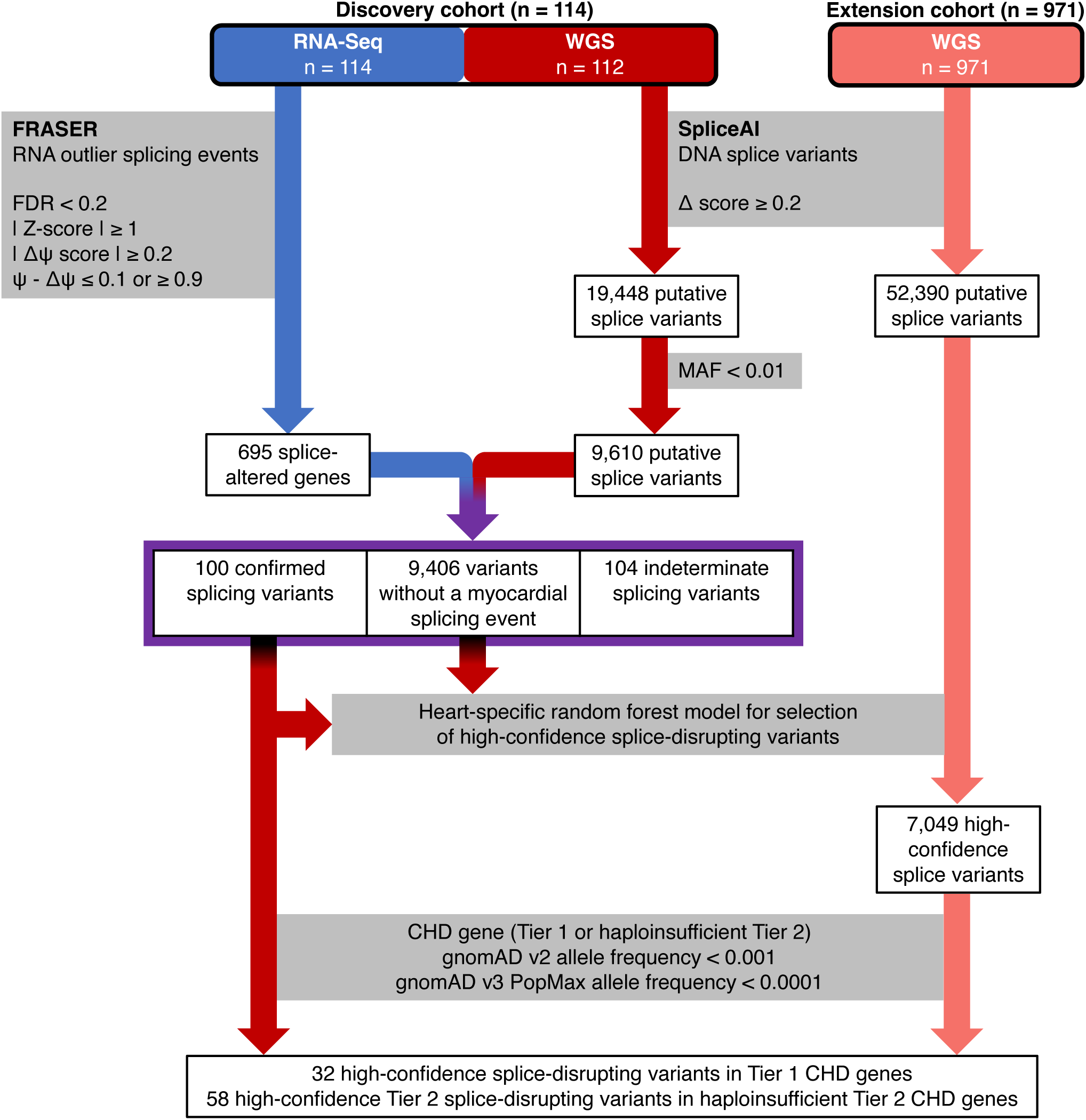
Schematic representation of the bioinformatics workflow to select high-confidence splice-disrupting variants in the Discovery and Extension cohorts. Variant selection strategy is shown. The heart-specific random forest model was independently validated on a set of 43 cardiomyopathy cases. This strategy ultimately yielded 32 high-confidence Tier 1 variants and 58 high-confidence Tier 2 variants. RNA-Seq, RNA sequencing; WGS, whole genome sequencing; FDR, false discovery rate; MAF, minor allele frequency; CHD, congenital heart disease

### Random forest model to predict cardiac relevant non-canonical splice-disrupting variants

Utilizing features from the set of confirmed splicing events, we trained random forest models to predict whether variants identified in WGS data are associated with aberrant splicing in human myocardium. Model 1 included only SpliceAI Δ scores as input. While SpliceAI has been reported to have good accuracy for detecting splice-disrupting variants, it utilizes only the genomic sequence of pre-mRNA transcript as input, which does not take into account existing splice junction boundaries or the likelihood of false positive variant calls. We therefore trained a second model 2 which included not only SpliceAI delta scores but also the variant distance to the nearest annotated splice junction, the variant type (SNV or indel), and whether the variant occurred in a branchpoint region, low complexity region, and/or repetitive region. As these two models include only DNA variant features, they are not trained to predict organ-specific splicing validation that may be unique to cardiac-expressed genes. A third model 3 was thus trained and included all of the aforementioned DNA variant features in addition to the corresponding median gene expression TPM value in patient myocardial samples. Due to the imbalance between the number of confirmed vs unconfirmed putatively splice-disrupting variants, all models were weighted to prioritize the selection of the confirmed class. The performance of all three models was internally assessed using five-fold cross validation. While all three models performed better than random, model 3 that included DNA variant and gene expression features provided highest performance accuracy (AUC=0.92 with five-fold cross-validation compared to 0.66 for model 1 and 0.82 for model 2) (**Figure 2**). Although model 3 prioritized gene expression values, SpliceAI Δ scores and distance from the nearest existing annotated splice site as the features of highest importance, other aforementioned features also provided independent predictive information. Together, this suggests that location and type of DNA variants and the cardiac expression of the genes in which they are found can improve the selection of cardiac-relevant high-confidence splice disrupting variants.

**Figure 2.**
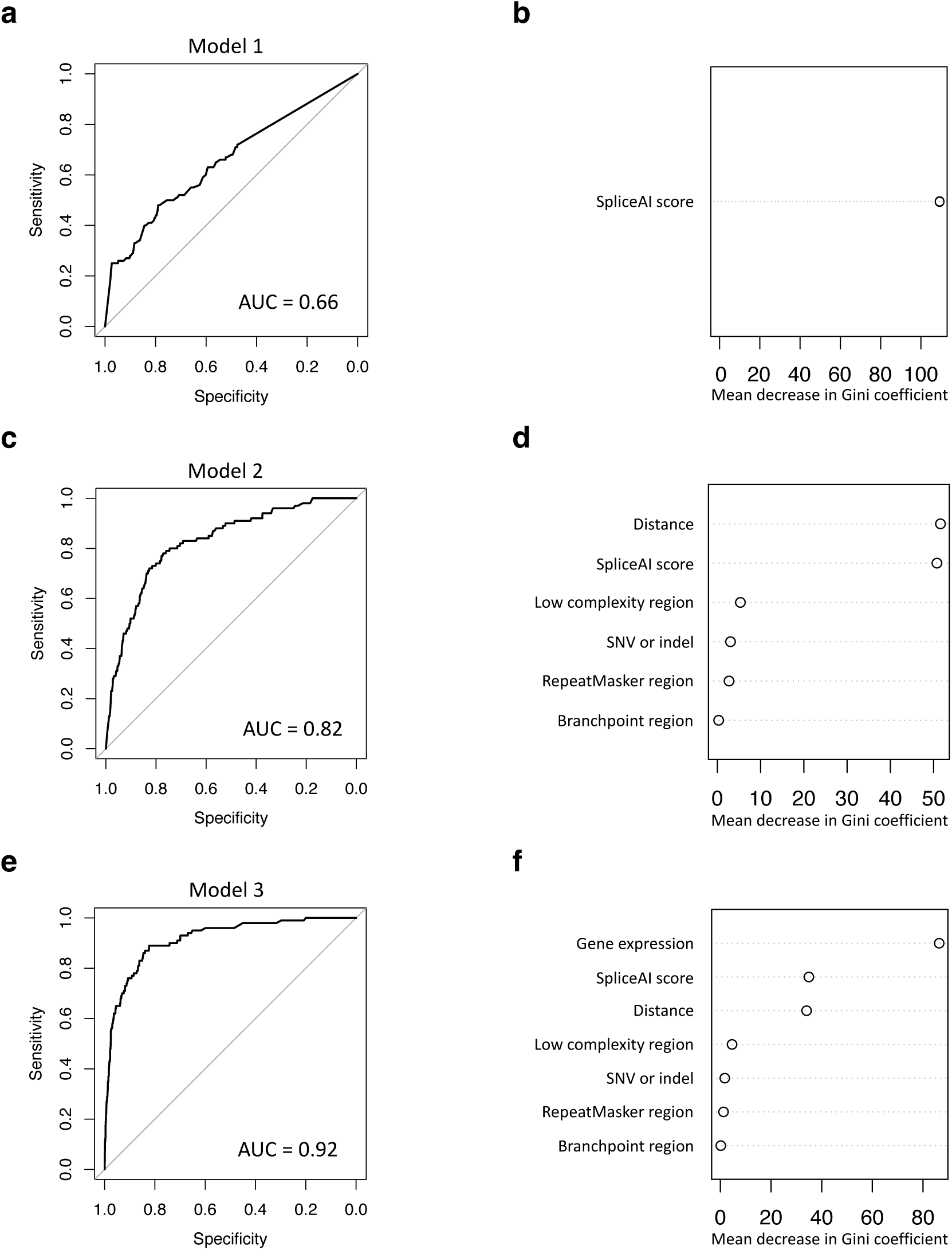
Performance of random forest models. Three types of models were designed and internal performance was assessed using five-fold cross-validation. Model 1 (a) and (b) used only SpliceAI scores, the Model 2 (c) and (d) used additional DNA variant features, and Model 3 (e) and (f) used all DNA features in addition to corresponding gene expression. The Gini coefficient is a measure for the probability that a specific feature is classified incorrectly when selected randomly. Area under the curves (AUC) are shown in (a), (c), and (e), while the relative importance of individual features are shown in (b), (d), and (f).

### Independent validation of the random forest model in a cardiomyopathy cohort

In order to externally validate variant selection by our random forest model, we investigated splice disrupting DNA variants in an independent cohort of 43 cardiomyopathy probands for whom both WGS and matching RNA-Seq profiles derived from the left ventricular myocardium were available^14^. We first selected SNVs and indels that were predicted with high sensitivity to result in the loss of a wild-type splice junction and/or the gain of a novel cryptic splice site (SpliceAI Δ score ≥ 0.2), and that were rare within this *Validation* cohort i.e. found in no more than one sample (MAF < 0.03). This identified 4,273 DNA variants across all genes, of which 529 were selected by the random forest model 3. We next identified aberrant splicing events in the *Validation* cohort using FRASER. Using the previous thresholds (false discovery rate < 0.2, an absolute Z-score ≥ 1, an absolute Δψ/θ score ≥ 0.2, and ψ/θ – Δψ/θ ≤ 0.1 or ≥ 0.9) yielded 360 observations of significantly-altered splicing events with a high effect size. We applied our random forest models to these data and showed that model 3 outperformed the other two models, with an area under the curve of 0.88 compared to 0.64 for model 1 and 0.76 for model 2 (**Figure 3**). Confirmed splicing variants, as well as all variants selected by model 3, are included in **Table 4**. While the smaller size of the *Validation* cohort limited the statistical power to identify outlier splice disruption events within the cohort, the absolute Z-scores of the altered splicing events was significantly higher in events harboring matching DNA variants selected by model 3 (p = 0.00041). We additionally investigated whether selected splice-disrupting DNA variants were associated with a reduction in gene expression, which may occur due to nonsense-mediated decay. Applying OUTRIDER^24^ to the *Validation* cohort, we observed that DNA variants selected by model 3 were associated with significantly reduced gene expression compared with variants that were not selected by the random forest model (p = 0.00076, mean Z-score = -0.258 vs -0.061). Of note, model 3 selected a pathogenic canonical splice site variant in a known cardiomyopathy gene, *FLNC*, previously reported by our group to be associated with reduced mRNA expression despite only a few RNA-Seq reads displaying abnormal splicing (presumably as a result of nonsense-mediated decay)^14^. Together these results confirm that our cardiac-specific random forest model improves the selection of putatively splice-disrupting variants in patients with childhood onset heart disease.

**Figure 3.**
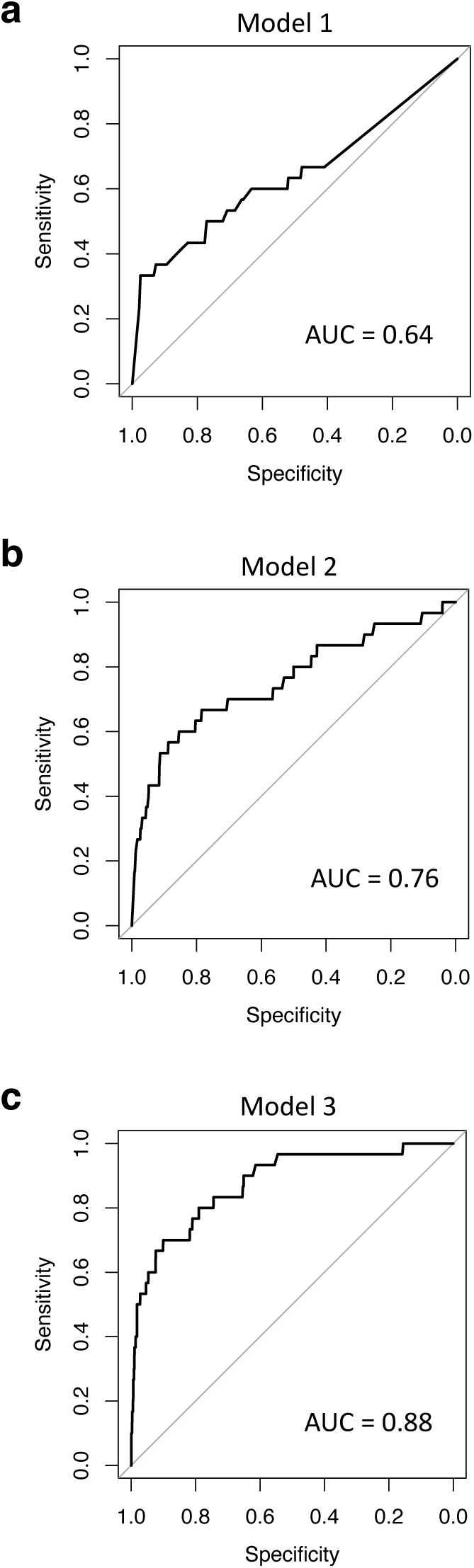
Model performance and examples of high confidence splice-disrupting events in the cardiomyopathy *Validation* cohort (n = 43). The random forest models were validated in with Area under the curves (AUC) ranging from 0.64 to 0.88 (a-c).

### High-confidence splice-disrupting variants in CHD genes in the *Extension* cohort

*Splice-disrupting variants in Tier 1 CHD genes*: In addition to the splice-disrupting variants identified in the *Discovery* cohort, we applied random forest model 3 to identify high-confidence splice-disrupting variants (SNVs and indels) in the *Extension* cohort of 971 patients with CHD (742 TOF, 229 TGA) with only WGS data. A total of 52,390 putative splice-disrupting variants were identified genome-wide (SpliceAI Δ score ≥ 0.2), of which model 3 selected 7,049 variants. Filtering for variants that were rare in controls (gnomAD v2 allele frequency < 0.001 and gnomAD v3 PopMax allele frequency < 0.0001) yielded 4,529 variants in 2875 genes. These genes were enriched for a diverse set of Human Phenotype Ontology terms that included abnormal heart and muscle morphologies (**Figure 4**, **Table 5**). We further narrowed our variant selection by limiting to those in a Tier 1 CHD gene with either a dominant mode of inheritance or homozygous in a gene with a recessive mode of inheritance. In total, we identified 32 high confidence splice-disrupting variants in 35 probands involving 22 Tier 1 genes (**Figure 5**, **Table 6**). Four variants were located at a canonical splice site, all of which were considered to be pathogenic/likely pathogenic by ACMG/AMP criteria. This included a splice donor variant in *TBX20*, which was validated by RNA-Seq (**Figure 6**). Intronic variants more than 10bp from existing splice sites accounted for 44% of high confidence Tier 1 splice-disrupting variants, many of which would not be detectable through panel or exome sequencing and would have been missed by applying only a high SpliceAI score cut-off. Of note, only one of these probands harbored additional pathogenic protein-coding variants in Tier 1 CHD genes to explain their CHD. None of these variants were observed in *Discovery* cohort samples, though two DNA variants in as many genes were observed in multiple unrelated probands in the *Extension* cohort.

**Figure 4.**
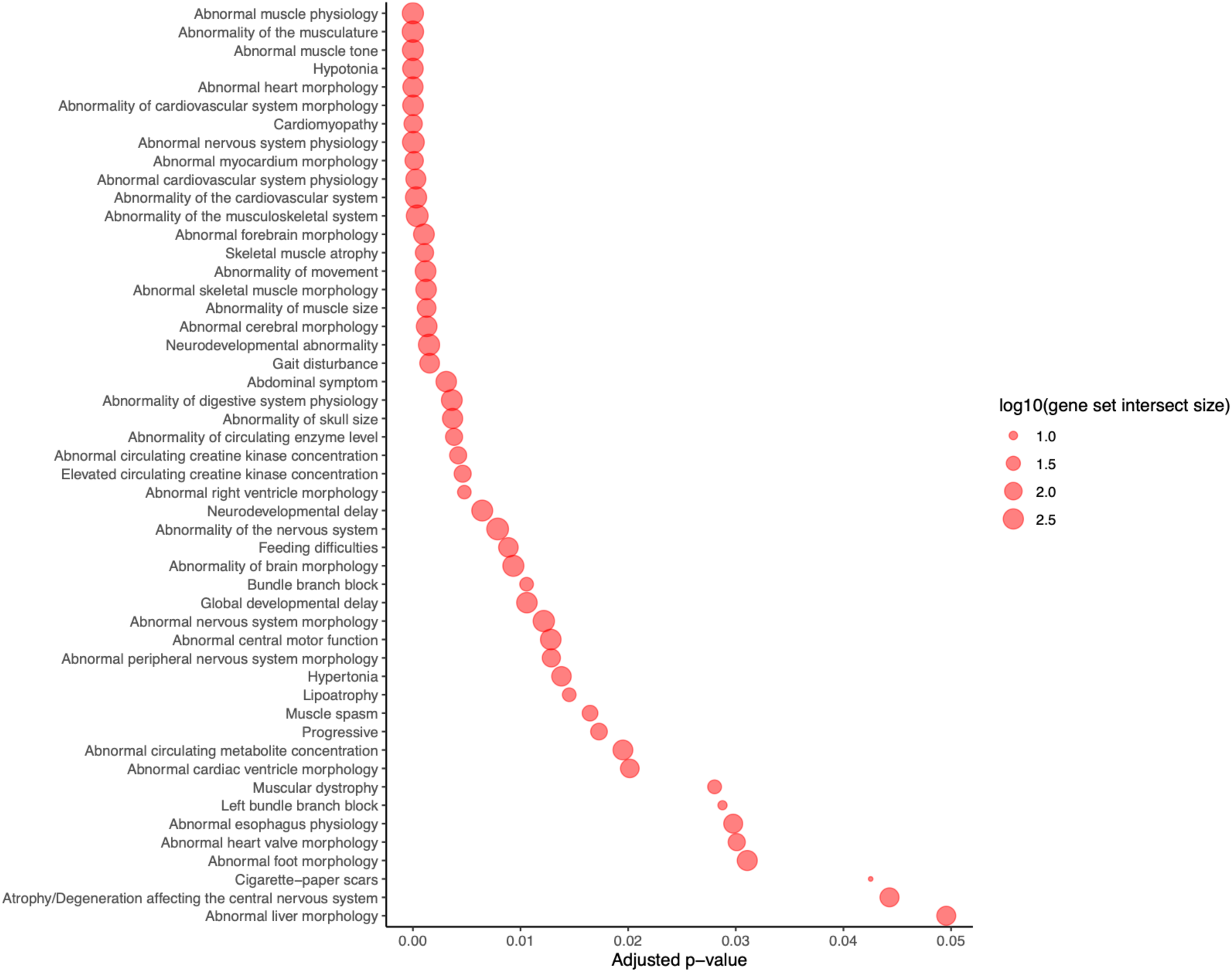
Significantly enriched gene sets in the *Extension* cohort (n = 971). Significantly enriched Human Phenotype Ontology (HP) terms among genes affected by high-confidence splice-disrupting variants selected by model 3.

**Figure 5.**
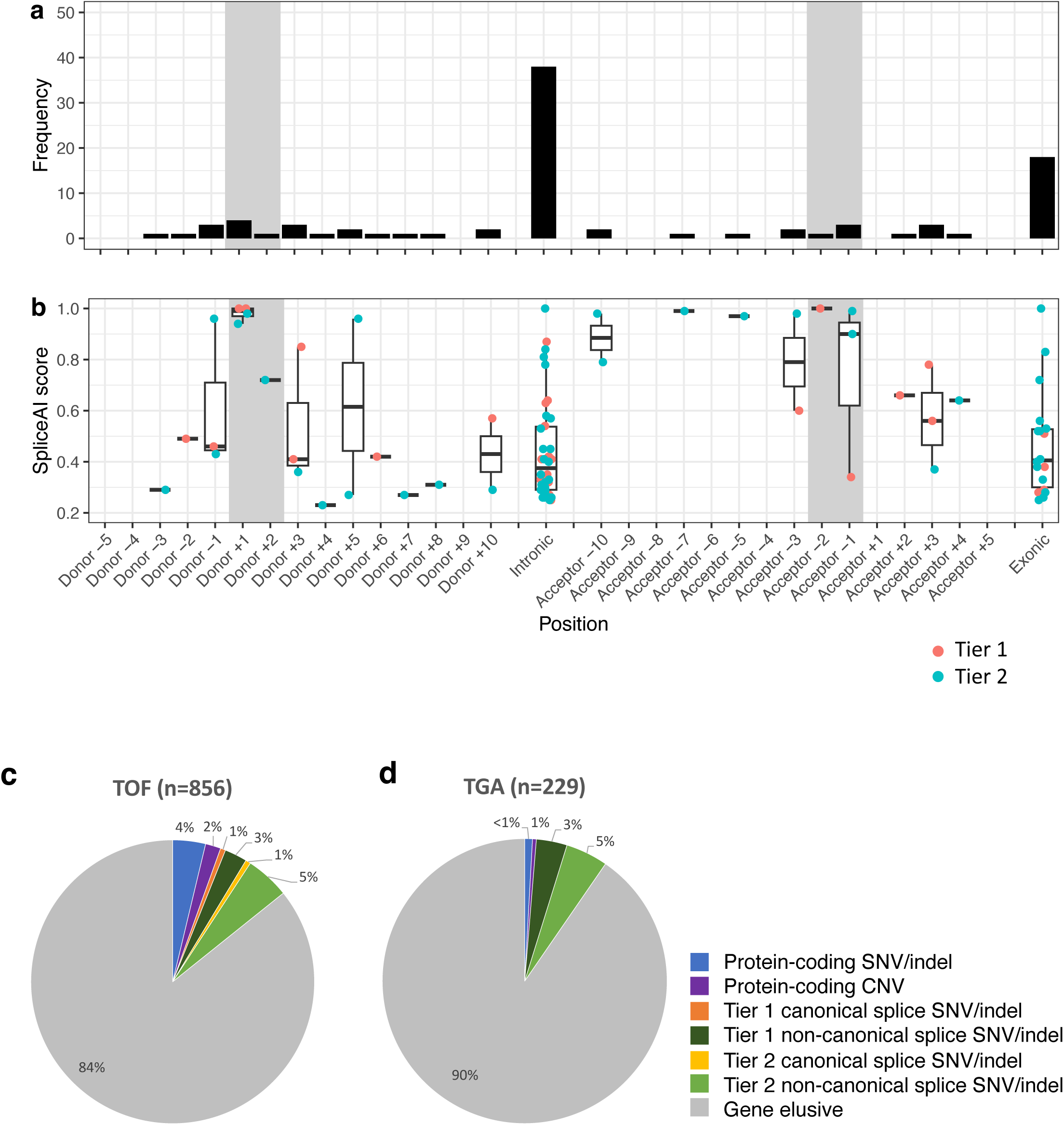
Summary of high-confidence splice-disrupting variant positions in CHD genes. 90 high confidence splice disrupting variants in CHD genes were identified in the Discovery (n=114) and Extension (n=971) cohorts. Variants were mapped to their closest annotated wild-type splice site within their corresponding gene. Canonical splice regions are highlighted in grey. (a) The frequency of variants with respect to relative positions. Intronic variants >10bp from a splice junction accounted for 41% of all variants. (b) SpliceAI Δ scores of variants. Our heart-specific model 3 selected variants with a large range of scores. Overall findings of pathogenic protein-coding variants and splice-disrupting variants in CHD genes are summarized for (c) TOF and (d) TGA samples across both the *Discovery* and *Extension* cohorts. TOF, tetralogy of fallot; TGA, Transposition of the great arteries; SNV, single nucleotide variant; indel, insertion-deletion

**Figure 6.**
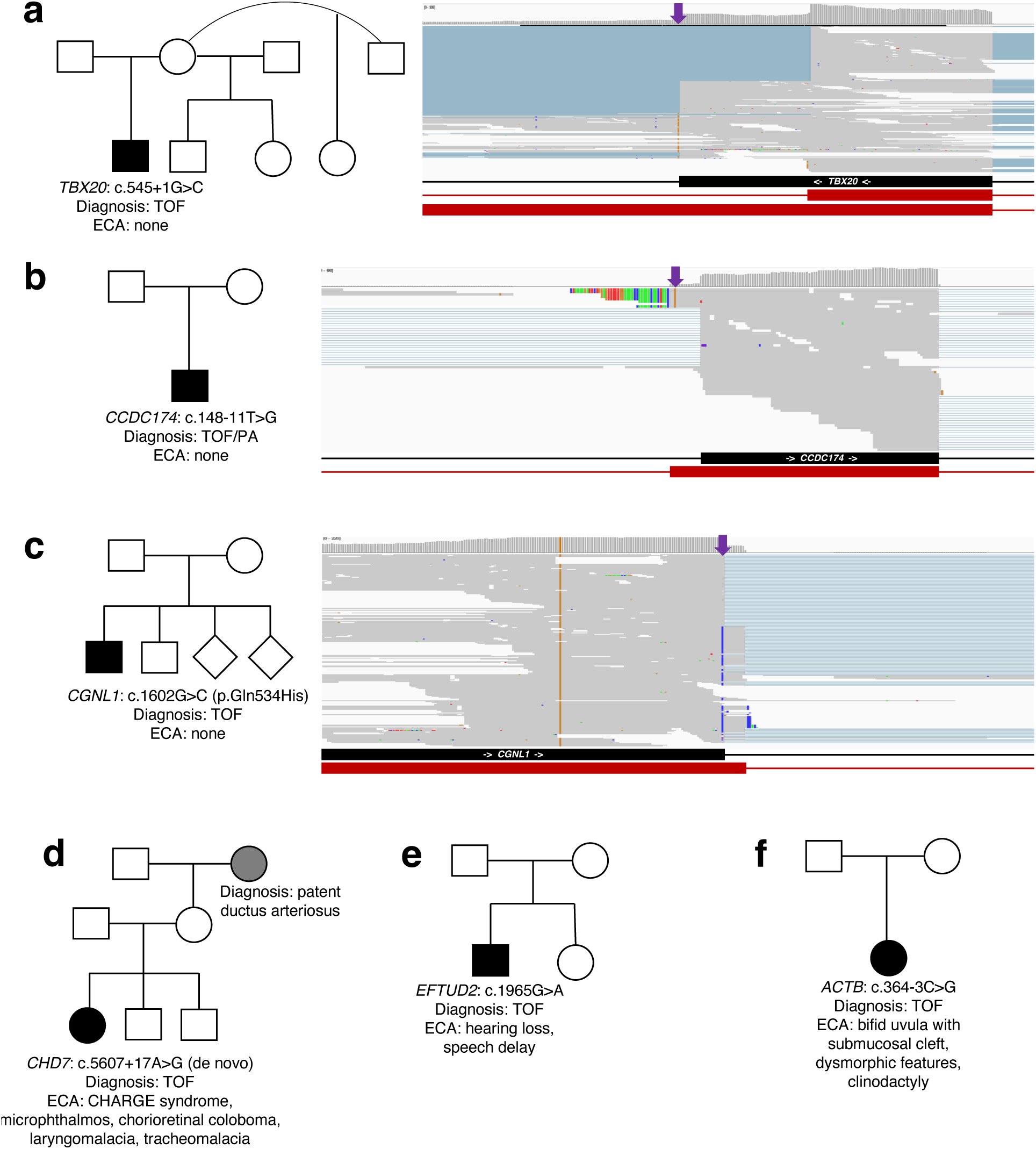
High confidence splice-disrupting variants in CHD genes. Family pedigrees and aberrant splicing events were observed in example Tier 1 genes (a) TBX20 (*Discovery* cohort), (d) CHD7 (*Extension* cohort), and (e) EFTUD2 (*Extension* cohort), as well as the Tier 2 genes (b) CCDC174 (*Discovery* cohort), (c) CGNL1 (*Discovery* cohort), and (e) ACTB (*Extension* cohort). Wild-type exon/intron boundaries below IGV screenshot are represented in black, and alternatively observed boundaries are represented in red. Arrows next to gene names represent reading direction. Purple arrows represent location of DNA splice-disrupting variant. TOF, tetralogy of fallot; ECA, extra cardiac anomalies.

#### Splice-disrupting variants in Tier 2 CHD genes

In addition to splice-disrupting variants in Tier 1 CHD genes, we searched for high-confidence splice-disrupting variants in the *Discovery* and *Extension* cohorts, that were rare (gnomAD v2 allele frequency < 0.001 and gnomAD v3 PopMax allele frequency < 0.0001) and occurred in haploinsufficiency intolerant Tier 2 CHD genes (gnomAD v2 pLI ≥ 0.9). This search yielded 58 variants in 34 genes among 61 probands (**Table 6**). Two of these variants were selected by model 3 in the *Extension* cohort and also observed in *Discovery* cohort samples, although neither appeared to validate with either aberrantly spliced RNA-Seq reads or reduced gene expression (Z-scores = -1.06 and 0.97). One pathogenic canonical splice site variant in the Tier 2 gene *TBX1* (c.867+1G>A) was observed in WGS data but not in RNA-Seq data from the same patient. This gene had low expression within the *Discovery* cohort (TPM = 0.06), and further investigation revealed that no RNA-Seq reads were aligned to this splice donor site. This is consistent with data suggesting that TBX1 is primarily expressed in the developing rather than adult heart^25^. The relative positions of these 58 variants were also consistent with what was observed in Tier 1 CHD genes, whereby deeply intronic variants (>10bp from an existing splice site) accounted for 40% of all Tier 2 variants. For 48 of 58 variants, the most probable predicted effect was the creation of a new cryptic splice site rather than the loss of an existing splice junction.

#### Genotype-phenotype correlation

We performed reverse phenotyping in probands harboring splice-disrupting variants in syndromic genes and identified some patients that harbored extra-cardiac features consistent with a syndrome even though it had not been clinically diagnosed at the time of the study. For example, one patient had an intronic *CHD7* c.5607+17A>G variant that was *de novo* with the patient demonstrating features consistent with CHARGE syndrome (**Figure 6**). This genetic finding triggered repeat genetic evaluation that led to confirmation of the diagnosis of CHARGE syndrome. A proband with classic TOF harboring a cryptic splice acceptor gain in the protein-coding region of *EFTUD2* also displayed a delay in speech, and mild to moderate hearing loss, likely of middle ear function, which required clinical intervention. One TOF proband harboring a splice region variant in the Tier 2 gene *ACTB* (c.364-3C>G) had extracardiac anomalies including bifid uvula with submucosal cleft, dysmorphic features, and clinodactyly, consistent with expected phenotype associated with defect in this gene.

One proband was observed to harbor intron retention in RNA-Seq for the *KRAS* gene, despite no candidate DNA variant being identified (**Figure 7**). This individual was found to have congenital malformations of the brain posterior fossa on reverse phenotyping, which may be consistent with a report of an activating *KRAS* mutation in a posterior fossa pilocytic astrocytoma^26^, and supports the pathogenicity of this alternative splicing event. An additional five significant splicing events in CHD genes were found without corresponding DNA variants (**Figure 7**, **Table 3**).

**Figure 7.**
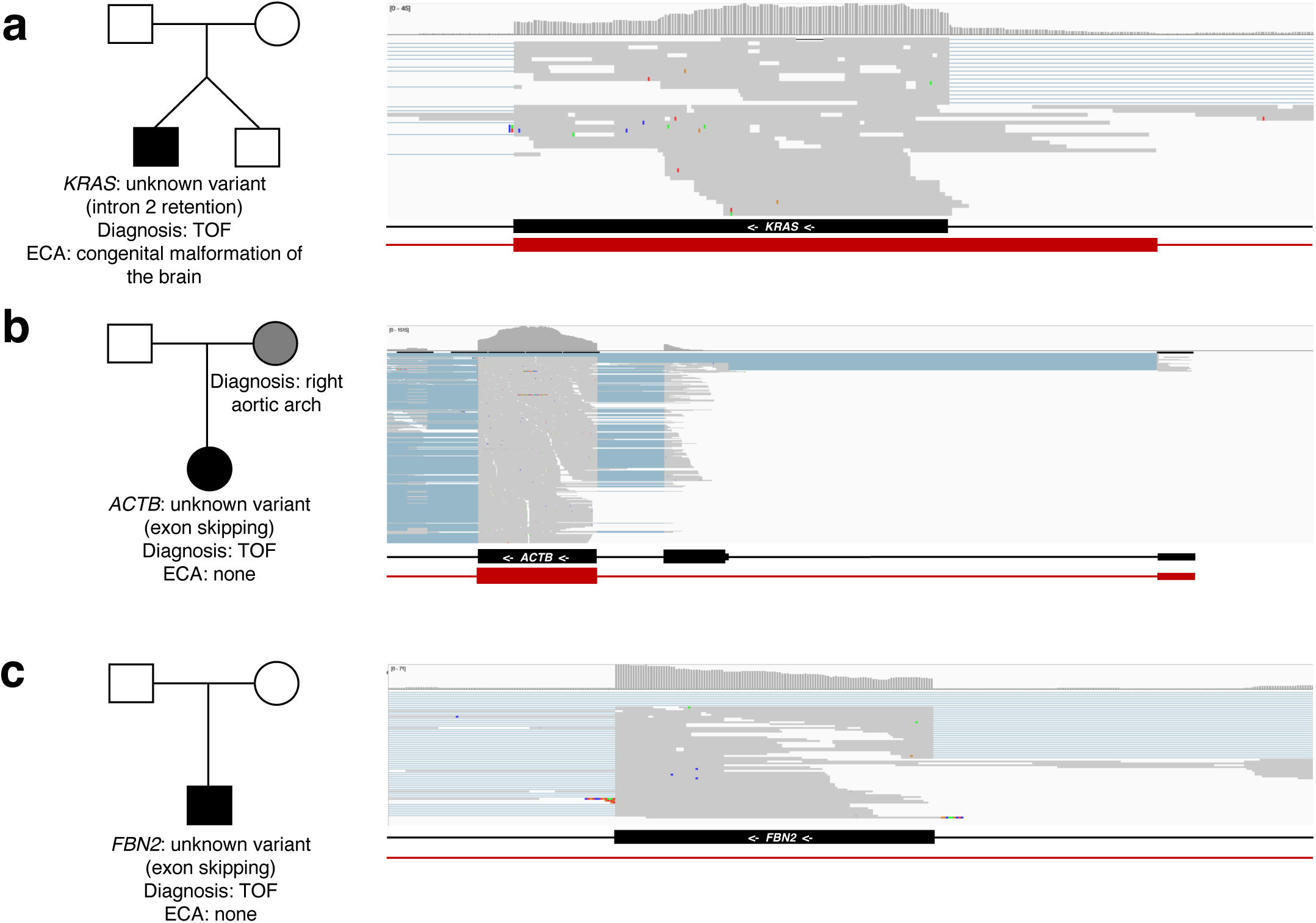
Significantly altered splicing occurring in CHD genes without an identified DNA variant. Family pedigrees and aberrant splicing events were observed in the *Discovery* cohort in CHD gene examples including (a) *KRAS*, (b) *ACTB*, and (c) *FBN2*. TOF, tetralogy of fallot; ECA, extra cardiac anomalies.

### Genome-wide burden of splice-disrupting variants in cases versus controls

We compared the characteristics of splice-disrupting variants in CHD patients to controls without CHD utilizing case-control burden analyses. We assessed the burden of rare (internal MAF < 0.01, gnomAD v2 allele frequency < 0.0001, and gnomAD v3 PopMax allele frequency < 0.0001), splice-disrupting (SpliceAI Δ score ≥ 0.2) high-confidence variants selected by our random forest model 3, in 971 *Extension* cohort CHD cases vs 2,570 healthy controls with WGS. The use of a stringent gnomAD PopMax threshold ensured that differences in cohort ancestries did not drive differences in burden between cohorts. Moreover, both gnomAD v2 and v3 thresholds were used in order to account for differences in variant frequencies of reference genomes between the *Extension* and *Control* cohorts.

Genome-wide, we did not observe a significant difference (p > 0.05) in variant burden in cases versus controls (**Figure 8**). However, there was a significantly higher burden in cases versus controls of splice-disrupting variants in all Tier 1-2 CHD genes (p = 0.011). On subgroup analysis, cases had a higher frequency of variants located at canonical splice sites, splice regions, protein-coding regions, and intronic regions of Tier 1 CHD genes, but these differences did not reach statistical significance. This may be due in part to the small variant numbers within gene sub-regions. While the *Extension* and *Control* cohorts had a similar proportion of male participants (57% and 49%, respectively), there was a lower proportion of samples of European descent in cases compared with controls (78% and 96%, respectively). To eliminate confounding related to ancestral differences between cases and controls, we further limited burden testing to individuals of European descent and observed similar trends, with a significantly higher burden of high-confidence splice-disrupting variants among CHD genes despite a reduced statistical power from the smaller sample size (p = 0.018).

**Figure 8.**
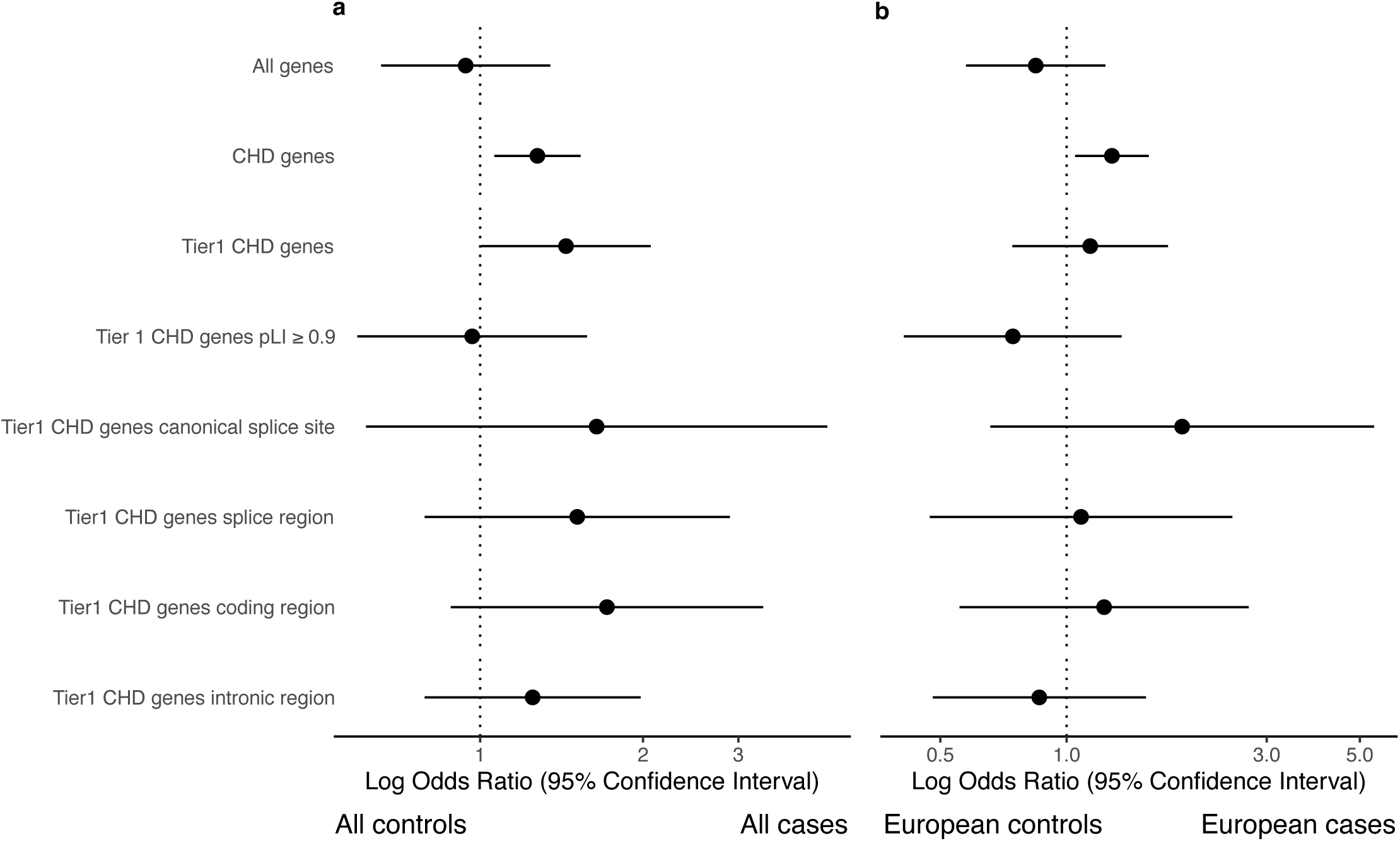
Case-control burden of variants. Variants were filtered and selected using increasingly conservative thresholds. Odds ratios and 95% confidence intervals are shown comparing n=971 CHD cases vs 2,570 healthy controls. (a) All samples. (b) Only samples of European descent. CHD, congenital heart disease; pLI, probability of being loss-of-function intolerant

In summary, a heart-specific model identified high-confidence splice-disrupting variants in CHD genes at canonical splice sites in 1% all CHD cases and non-canonical variants in 8% cases, with splice-disrupting variants accounting for 58% of putatively disease-causing variants in CHD cases (**Figure 5**). In particular, deeply intronic cryptic splice variants represented 41% of all DNA splicing variants in CHD genes.

## DISCUSSION

Our study applied whole genome sequencing coupled with myocardial RNA-Seq to identify and validate non-canonical splice-disrupting variants associated with CHD. Non-canonical splice-disrupting variants in coding regions were enriched in cases compared to controls and were associated with altered myocardial gene splicing. The findings were leveraged to develop a machine learning model to predict cardiac-specific, high-confidence non-canonical splice variants. Together, this first of a kind model identified disease-associated splice-disrupting variants in 9% of CHD patients (58% of all pathogenic CHD associated variants), representing a significant expansion of the role of these variants in heart disease.

RNA-Seq of patient myocardium enabled us to directly identify splice-disrupting events across the genomes of 114 TOF probands. By integrating these validated events with DNA variants obtained from WGS, we generated an *in silico* model that accurately predicted heart-specific splice-disrupting variants with greater accuracy than using DNA variant features alone. Application of the model to 971 CHD WGS samples, in addition to the confirmed splice variants assessed in patient myocardium of 114 cases, identified 3% of CHD cases that harbor non-canonical splice-disrupting variants in Tier 1 CHD genes. As nearly half (44%) of these variants were intronic, our combined use of RNA-Seq and WGS enabled the identification of putatively pathogenic variants that would not be detected by panel or exome sequencing, as intronic variants beyond ∼50bp are not reliably found with these methods^12,13^.

A recent whole exome sequencing study highlighted a role for splice-disrupting variants in CHD^9^. In particular, 2% of genome-wide, de novo, computationally-predicted splice-disrupting variants in CHD probands were validated by minigene assays. In addition, in a case-control burden analysis, an enrichment for rare splice region variants predicted to result in the loss of nearby existing splice junctions among CHD genes was observed. Unfortunately, exome sequencing is unable to detect deeply intronic splice-disrupting variants^12,13^, and minigene assays alone have technical limitations since they are not cardiac specific, cannot test variants in repetitive regions, and often provide indeterminate results^27,28^.

In this regard, our machine learning model was highly accurate for identifying non-canonical variants that result in confirmed splicing events specific to the human heart (AUC=0.92). When applied to a cohort with cardiomyopathy, the model was able to identify non-canonical variants that affect splicing in cardiomyopathy genes. This approach allowed us to recover deeply intronic cryptic splicing variants that cannot be captured by whole exome sequencing, and have not been previously reported. Of note, direct investigation of patient myocardium identified aberrant splicing events in CHD and other cardiac genes even when a causal DNA variant responsible for the effect could not be definitively confirmed. This may be due to the causal variant having a predicted SpliceAI Δ score below our minimum threshold (0.2), the variant affecting splicing at a greater distance than our maximum threshold (100bp), or somatic mosaicism resulting in variants in the heart that are undetectable in blood and/or saliva^29^.

81% of predicted high-confidence Tier 1 CHD splice-disrupting variants occurred in genes with associations to syndromic disease that had not been clinically identified. Intriguingly, an intronic *de novo* variant in *CHD7* was observed in a TOF proband who was negative on clinical genetic testing, and had no family history of heart disease (**Figure 6**). Pathogenic variants in *CHD7* are associated with CHARGE syndrome, and this patient had phenotypic features consistent with this disease, supporting the pathogenicity of the detected high-confidence cryptic splicing variant. Crucially, a prediction model trained only on SpliceAI scores failed to select this variant, again reinforcing the value of using a heart-specific prediction model. Additional splice-disrupting events were observed in the syndromic genes *EFTUD2* and *KRAS* (**Figure 7**), with patient phenotypes consistent with known genotype-phenotype associations.

While at this time, most non-canonical splice-disrupting variants must be functionally validated in order to be considered pathogenic/likely pathogenic, our use of variant segregation and deep phenotyping meant that we were able to classify variants like the aforementioned *CHD7 de novo* variant as likely pathogenic. As computational tools continue to improve the accuracy of variant selection with specific effects on splicing, it may allow for more streamlined clinical reporting of such variants. Indeed, our results support and extend on the recently published ACMG/AMP framework for validating and reporting splice-disrupting variants, including those outside of canonical splice sites^11^. In particular, our findings reinforce the utility of using heart-specific models trained on patient myocardium to improve the accuracy of variant selection in CHD. Moreover, our observation that deeply intronic cryptic splice variants contribute to CHD highlight the necessity of including these types of variants in clinical tests.

Our study had some limitations. While RNA-Seq of patient myocardium allowed us to directly identify altered splicing events *in vivo*, these types of events may sometimes lead to nonsense-mediated decay, thereby limiting the ability to detect them in patient tissue. As none of the *Discovery* or *Validation* cohort patients were treated with compounds to inhibit nonsense-mediated decay prior to resection of their myocardial tissue, some variants affecting splicing may therefore have been classified as having an unconfirmed splicing effect. Within our *Discovery* cohort, 22% of CHD genes had very low expression (TPM < 1). Similarly, CHD genes expressed during embryogenesis but not in mature patient myocardium may not have been detectable in our RNA-Seq data. While *in vitro* methods for the inhibition of nonsense-medicated decay can be used to validate splicing effects, they are not feasible to test for a large number of variants. Another limitation is that short-read sequencing for both RNA-Seq and WGS may have missed variants in homologous and low complexity regions, due to unreliable alignments in such regions. Similar studies performed with higher depth sequencing and/or long-read sequencing may further extend our ability to reliably detect splice-disrupting events in known and candidate CHD genes. Finally, while we attempted to control for differences in alignment and variant calling within our case-control burden analysis, we acknowledge the confounding effect of these differences when conducting burden analyses across different cohorts.

In summary, our findings that non-canonical splice-disrupting variants contribute to the genetic etiology of CHD make a strong case for routine evaluation of intronic variants as well as to assess the splicing effect of variants in protein-coding regions. Our cardiac-specific bioinformatic model that utilizes both RNA-Seq and WGS-based variant annotation predicted with high confidence splice-disrupting variants that make a significant contribution to the genetic basis of CHD.

## METHODS

### Study Cohorts

#### Congenital heart disease (CHD) cases

The overall cohort included 1085 probands, of which 856 had TOF and 229 had TGA (**Table 1**). Among these cases, 479 TOF and 229 TGA were enrolled through the Heart Centre Biobank Registry at the Hospital for Sick Children (Ontario, Canada), 250 TOF were enrolled through the Kids Heart BioBank at the Heart Centre for Children, The Children’s Hospital at Westmead (Sydney, Australia), and 127 TOF were enrolled through the CONCOR registry at the Amsterdam Medical Center (Netherlands).

The *Discovery cohort* included 114 unrelated tetralogy of Fallot (TOF) from Ontario. All probands had myocardium available; blood or saliva was also available for 112/114 probands.

The *Extension* cohort included 971 unrelated TOF and TGA probands and 191 family members enrolled in the Heart Centre Biobank Registry from Ontario; the Kids Heart BioBank at the Heart Centre for Children, The Children’s Hospital at Westmead (Sydney, Australia); and the nation-wide Dutch CONCOR registry at the Amsterdam Medical Center (Amsterdam, Netherlands). Blood or saliva derived DNA was available for all probands.

The *Validation* cohort included 43 unrelated cardiomyopathy probands enrolled in the Heart Centre Biobank Registry at the Hospital for Sick Children (Ontario, Canada). All probands had myocardium, and blood or saliva, available.

Collection and use of biospecimens through the registries was approved by local or central Research Ethics Boards and written informed consent was obtained from all patients and/or their parents/legal guardians and study protocols adhered to the Declaration of Helsinki.

#### Controls

The control cohort included 2,570 whole genome sequencing (WGS) samples from the Medical Genome Reference Bank (MGRB)^15^. MGRB variants were obtained from the original publication, after alignment to GRCh37 and variant calling for all samples. Single nucleotide variants (SNVs) and insertion-deletions (indels) were converted to hg38 using LiftoverVcf (http://broadinstitute.github.io/picard). *Control* cohort characteristics are provided in **Table 1**.

### Whole genome sequencing

WGS of CHD and cardiomyopathy cases was performed on high quality DNA from blood or saliva of probands and their family members using the Illumina HiSeq X or NovaSeq platform by The Centre for Applied Genomics (TCAG, The Hospital for Sick Children, Toronto), or Macrogen (South Korea). Illumina TruSeq DNA PCR-Free kits were used for library preparation. High-quality paired-end reads (2×150bp) were mapped to human genome reference sequence hg38 using one of two workflows as follows:

Paired-end raw reads were trimmed and cleaned by trimmomatic v.0.32^30^, then mapped to human reference genome hg38 using bwa v.0.7.15^31^. The reference genome sequence and training datasets were downloaded from the Genome Analysis Toolkit (GATK) resource bundle (ftp.broadinstitute.org/bundle)^32^. Mapped reads were realigned and calibrated by base quality score recalibration tools (GATK v4.1.2.0). HaplotypeCaller was used to generate genotype Variant Call Format (gVCF) files for each sample, then gVCF files for batches of samples were combined and joint-called by using CombineGVCFs and GenotypeGVCFs tools. In order to filter out probable artifacts in the calls, SNVs and indels were recalibrated separately by variant quality score recalibration (VQSR) tools, and variants that passed VQSR truth sensitivity level 99.5 for SNPs and level 99.0 for indels were retained. The VariantFiltration tool was used to mark out the low Genotype Quality (GQ) SNV and indel sites whose GQ values were lower than 20 and read depths were lower than 10. Copy number variants (CNVs) were called as described in ^33^, using ERDS^34^ and CNVnator^35^. Structural variants (SVs) were called using Manta^36^ and Delly^37^. Sample ancestry and relatedness among family members was estimated and verified using somalier v0.2.11^38^ with default parameters.

WGS data for the MGRB control cohort were generated as previously reported^15^. Briefly, DNA was extracted from blood and Illumina TruSeq Nano DNA High Throughput kits were used for library preparation. Reads were sequenced on Illumina HiSeq X, then aligned to the 1000 Genomes Phase 3 decoyed version of build 37 of the human genome using GATK best practices. GATK HaplotypeCaller was used to generate gVCFs for SNVs and indels, then joint-called in a single batch using GATK GenotypeGVCFs.

### CHD gene list

Tier 1 CHD genes were selected based on a moderate, strong, or definitive association with CHD according to ClinGen criteria^16^. We further annotated and categorized additional CHD genes using (1) published literature; (2) existing databases including Online Mendelian Inheritance in Man (OMIM)^17^, Clinical Genome Resource (ClinGen)^18^, and CHDgene^19^; (3) inclusion in clinical gene panels; and (4) expert curation and classified genes with a limited evidence for association with CHD as Tier 2 genes. Canonical transcriptional isoforms were annotated using Matched Annotation from NCBI and EMBL-EBI (MANE)^39^. Gene constraint annotations were obtained from gnomAD (v2)^40^ (**Table 2**).

### Interpretation of protein-coding and canonical splice site variants

Variant interpretation for pathogenicity was performed using American College of Medical Genetics and Genomics and the Association for Molecular Pathology (ACMG/AMP) criteria^20,21^. SNVs and indels were first annotated for pathogenicity using InterVar v2.0.2^41^. Variants with internal Human Gene Mutation Database (HGMD) Pro 2019^42^ classifications of Disease-associated polymorphism with supporting functional evidence (DFP) were assigned a PS3 score, while variants with an internal classification of Disease-associated polymorphism (DP) or Disease causing mutation (DM) were assigned a PP5 score. Variants classified by InterVar as “Pathogenic” or “Likely pathogenic” and occurring in Tier 1 or Tier 2 CHD genes were subsequently manually reviewed. Gene inheritance and associated disease conditions were obtained from OMIM^17^. Variants in recessive genes were required to either have a homozygous or bi-allelic genotype. Ratios of observed/expected (o/e) loss-of-function (LoF) or missense variants for affected genes were obtained from the Genome Aggregation Database (gnomAD) v2.1.1^40^. Population variant allele frequencies were obtained from gnomAD v3.1.2. Where possible, variant segregation among family members was considered. Variants in genes for dominant disorders had allele frequencies <0.01% for PM2, between 1% and 5% for BS1, and >5% for BA1.

Variants in genes for recessive disorders had allele frequencies <0.1% for PM2, between 2% and 10% for BS1, >10% for BA1. The UCSC Genome Browser was used to investigate low mappability and RepeatMasker annotations^43^. Variant reads were manually inspected using the Integrative Genomics Viewer (IGV)^44^ to exclude any likely false positive variants with insufficient evidence or insufficient read coverage. Variants with a heterozygous genotype call and a variant allele fraction of less than 33% or greater than 66%, variants with <20x coverage, and variants with many mismatched bases in nearby reads were excluded. ClinVar^45,46^ was used to search for any pre-existing classifications or other variants occurring at the same nucleotide or amino acid position.

For CNVs and SVs, automatic filters were applied and variants were retained if they met the following criteria: (1) are absent from or present at 1% frequency or less in a database of CNVs/SVs, generated from Illumina HiSeqX sequencing data of parents of children with autism spectrum disorder at TCAG^47^ and called by using the same methodology. (2) variants that overlapped with an exonic region, and (3) overlapped with a gene in the CHD genelist (with the exception of *de novo* variants which were assessed even if they did not overlap a CHD gene). To reduce the number of false positive CNVs, only variants called by both ERDS and CNVnator were retained. Each variant that passed these automatic filters was queried through the DECIPHER browser^48^. Variants that overlapped a substantial number of benign/population variants in DGV^49^ or gnomAD Structural Variants^50^ were not further considered, depending on the suspected mode of inheritance. Inversion breakpoints were queried through the UCSC genome browser^51^ – inversions whose breakpoints did not intersect CHD genes were not further considered. The remaining variants were visualized using either IGV^44^ or Samplot^52^, depending on their size and complexity, to confirm their authenticity. Variants that passed the above inspections proceeded to manual ACMG/AMP classification^21^. Intragenic CNVs/SVs were submitted to AutoPVS1^53^, to automatically assign a PVS1 criterion for haploinsufficient genes (i.e. genes with a pLI score greater than or equal to 0.9, a ClinGen dosage curation indicating haploinsufficiency, or literature evidence supporting a loss-of-function pathogenic mechanism). Complex variants (called as dual DUP-DEL, DUP-DEL-INV etc.) were only considered if there was phenotype support for the genes harboring them. We surveyed the literature and databases such as OMIM and ClinVar to identify similar CNVs/SVs reported in individuals with the phenotype of interest. Inheritance data if available was taken into account when classifying variants (whether the variant was de novo, parentally inherited, or unknown), as per the aforementioned ACMG/AMP guidelines. Visualization was used to confirm inheritance calls.

### Detection of putatively splice-disrupting variants in WGS data

Putatively splice-disrupting WGS SNVs and indels were identified using SpliceAI^22^. Exome annotations and splice junctions were similarly obtained from SpliceAI (https://github.com/Illumina/SpliceAI). Pre-computed masked SpliceAI delta scores were utilized where possible (https://basespace.illumina.com/projects/66029966), otherwise SpliceAI (v1.3.1) was used to generate masked delta scores with a maximum distance of 100bp between the variant and gained/lost splice site. Variants with a SpliceAI delta score ≥ 0.2 were retained and subsequently annotated with the predicted effect (VEP v102^54^), reported pathogenicity (ClinVar 2022-04-03 and HGMD Pro 2019), control allele frequency (gnomAD v3.1.2), gene constraint (gnomAD v2.1.1), genomic low complexity regions (https://github.com/lh3), genomic RepeatMasker regions (https://www.repeatmasker.org), and wild-type splicing branchpoints^55,56^. Ensembl RNA transcripts were further annotated as canonical by MANE v1.0 (MANE Select or MANE Plus Clinical)^39^.

### Myocardial RNA sequencing

RNA sequencing (RNA-Seq) was performed on right ventricular myocardial samples available from 114 unrelated TOF probands and 43 unrelated cardiomyopathy probands from SickKids Heart Centre Biobank. Myocardium was obtained from patients who had consented to biobanking from leftover tissue at the time of cardiac surgery and was immediately snap-frozen in the operating room and stored in liquid nitrogen. None of the patients received inhibitors of nonsense-mediated decay prior to tissue resection. Total RNA was extracted from myocardial samples using the RNeasy Mini kit (QIAGEN, Canada). RNA-Seq was performed using Illumina HiSeq 2500 or NovaSeq platforms at The Centre for Applied Genomics (TCAG, The Hospital for Sick Children, Toronto). Raw sequencing reads were trimmed by Trimmomatic v0.36^30^ for quality trimming and adapter clipping. The remaining reads were aligned to the GRCh37 reference genome (1000 Genomes Project reference genome, hs37d5) using STAR (v2.6.1.c)^57^ with basic two-pass mode and Ensembl GTF (release version 87)^58^ was used for the annotation. Gene and transcript expression level quantification were prepared using RSEM (v1.2.22)^59^.

### Identification of aberrant splicing events in myocardial RNA-Seq data

Aberrant splicing events were identified in RNA-Seq data using FRASER v1.8.1^23^. Introns with unreliable detection were filtered out using the ‘filterExpressionAndVariability’ method with default parameters except for requiring a minimal read count of 15 in at least one sample. An ‘AE’ denoising autoencoder with a BB loss was used to fit the splicing models, with hyperparameters ψ5=14, ψ3=14, and θ=7 for the *Discovery* cohort and ψ5=5, ψ3=4, and θ=2 for the *Validation* cohort. Optimal hyperparameters were determined using the ‘optimHyperParams’ method. Splice events were annotated using biomaRt as part of the ‘annotateRanges’ method^60^. Observed events were considered to be significant with a false discovery rate < 0.2, an absolute Z-score ≥ 1, an absolute Δψ/θ score ≥ 0.2, and ψ/θ - Δψ/θ ≤ 0.1 or ≥ 0.9. Events annotated as not mapping to a gene or to multiple genes were excluded. All reported splicing events in CHD genes were visually inspected; ambiguous and apparent false positives were excluded.

### Gene expression outlier analysis in myocardial RNA-Seq data

Gene expression outliers in RNA-Seq data were identified using OUTRIDER (v1.8.0)^24^. OUTRIDER was run on the *Discovery* (TOF) and *Validation* (CMP) cohorts separately. Low-expressed genes were first filtered out by selecting genes with at least 10 read counts in more than one-third of the input samples. Before fitting the input cohort to the OUTRIDER model, the optimal encoding dimension “q” was first determined by using the “findEncodingDim” method. The encoding dimension was estimated to be 19 for the TOF model and 8 for the CMP model. Genes with a false discovery rate < 0.2 were considered as significant expression outliers.

### Generation of random forest models for selecting heart-specific splice-disrupting variants

Genome-wide variants were classified by whether or not they were associated with confirmed splicing events by searching for matching significant events within ± 100bp of altered splicing boundaries called by FRASER. To test for the enrichment of univariate features that associate with variants that validated by FRASER, we used two-sided Mann-Whitney U tests for continuous variables and two-sided Fisher’s exact tests for binary variables. The R package randomForest^61^ was used to create and test three machine learning models for the prediction of splice-disrupting variants. Model 1 used only SpliceAI Δ scores as input; model 2 included SpliceAI delta scores in addition to the variant distance to the nearest annotated splice junction, the variant type (SNV or indel), and whether the variant occurred in a branchpoint region, low complexity region, and/or repetitive region; model 3 included all of the aforementioned DNA variant features in addition to the corresponding median gene expression TPM value in RNA-Seq data. Training classes included 100 confirmed DNA variants versus 9,406 DNA variants without a confirmed effect on splicing. Variants with missing feature values were omitted by the models (i.e. missing values were not imputed). To account for the imbalance in the training class frequencies, variant class was inversely weighted by the corresponding number of observations in the training data. All models were internally evaluated for performance (AUC) using five-fold cross-validation. The models were subsequently retrained on the entire *Discovery* cohort prior to its application to additional cohorts.

### Selection of high-confidence splice-disrupting variants using the optimal random forest model

The optimal random forest model 3 was applied to WGS data from the *Extension* and *Control* cohorts in order to predict high-confidence splice-disrupting variants. Variants were further filtered to include only those rare in gnomAD control populations (gnomAD v3 PopMax filtering allele frequency at 95% confidence < 0.0001), and visually inspected with likely false positive calls subsequently removed. For the case-control burden analysis, both gnomAD v2 and v3 statistics were used in order to account for different the reference genomes used to align the *Extension* (hg38) and *Control* (GRCh37) cohorts (gnomAD v2 exome allele frequency < 0.0001, gnomAD v2 genome allele frequency < 0.0001, and gnomAD v3 PopMax filtering allele frequency at 95% confidence < 0.0001). To further reduce possible false positive variant calls, variants were additionally limited to those that were rare and within both our *Extension* cohort (internal allele frequency < 0.01) and *Control* cohort (internal allele frequency < 0.01). P-values were calculated using a two-sided Fisher’s exact test. To reduce bias in odds ratios calculations and avoid “zero cells” in the contingency tables, 0.5 was added to each observed cell frequency (Haldane-Anscombe correction).

### Gene set analysis

Enrichment analysis of gene sets was performed using g:Profiler core tool g:GOSt with default parameters^62^. Genes were input as Ensembl identifiers. An adjusted p-value threshold of 0.05 was used to determine significance. Adjusted p-values were calculated using the g:SCS (Set Counts and Sizes) method which considers dependencies between multiple tests by taking into account the overlap in functional terms^63^.

### Data analyses and visualizations

All aforementioned statistical analyses, as well as data visualizations, were carried out using the R Programming Environment v4.1.2. Two-sided t-tests were used to compare the Z-scores of variants associated with RNA-Seq outlier splicing or expression events. Graphical data plots were created using the ggplot2^64^ and pROC^65^ libraries.

## Data Availability

Sequencing data for the Discovery and Extension cohorts will be deposited in the European Genome-Phenome Archive (EGA), and will be available for download upon approval by the Data Access Committee. Sequencing data for the cardiomyopathy Validation cohort is available in EGA under accession EGAS00001004929, and are available for download upon approval by the Data Access Committee. Control cohort MGRB data are available by controlled access in EGA under accession EGAS00001003511. Additional data generated or analyzed during this study are included in the supplementary information files, and additional raw data used for figures and results are available from the corresponding author on reasonable request.

https://ega-archive.org/studies/EGAS00001004929

https://ega-archive.org/studies/EGAS00001003511

## ACKNOWLEDGEMENTS

We acknowledge the Labatt Family Heart Centre Biobank at the Hospital for Sick Children for access to DNA and RNA samples, and The Centre for Applied Genomics at the Hospital for Sick Children for performing WGS and RNA-Seq. We thank Roderick Yao for his work in identifying and assessing putatively pathogenic protein-coding and canonical splice site SNVs and indels in CHD genes. We thank Doris Škorić-Milosavljević for valuable help regarding the patients obtained through the CONCOR registry. We also thank the patients and family members who participated in this study. The results shown here are in whole or part based upon data generated by the MGRB Partners: https://sgc.garvan.org.au/initiatives/mgrb. The Medical Genome Reference Bank was funded by the NSW State Government. This study makes use of data generated by the DECIPHER community. A full list of centres who contributed to the generation of the data is available from https://deciphergenomics.org/about/stats and via email from. DECIPHER is hosted by EMBL-EBI and funding for the DECIPHER project was provided by the Wellcome Trust [grant number WT223718/Z/21/Z].

## CONTRIBUTIONS

RL and SM conceived and designed the work, drafted the work and substantively revised it. SM acquired funding for the project. All authors contributed to acquisition, analysis or interpretation of data. All authors approved the submitted version and have agreed both to be personally accountable for the author’s own contributions and to ensure that questions related to the accuracy or integrity of any part of the work, even ones in which the author was not personally involved, are appropriately investigated, resolved, and the resolution documented in the literature.

